# Multimodal Phenotyping of Myofascial Pain Syndrome Using Rotational Shear Wave Elastography and Clinical Network Analysis

**DOI:** 10.64898/2026.07.23.26358787

**Authors:** Matin Jahani Jirsaraei, Yu-Lin Hsu, Reihana Akhwand, Abhishek Aher, Seiyon Lee, Secili DeStefano, John Srbely, Jay Shah, William Rosenberger, Samuel Acuña, Yonathan Assefa, Lynn H. Gerber, Siddhartha Sikdar

## Abstract

Myofascial pain syndrome (MPS) is characterized by increased muscle stiffness, trigger points, and functional limitations, yet clinical diagnosis remains largely subjective. Shear wave elastography (SWE) provides quantitative assessment of muscle mechanical properties, but its value for identifying biomechanical and clinical phenotypes of MPS is not fully established. This study evaluated whether stiffness parameters derived from multi-angle SWE can reliably characterize upper-trapezius anisotropy, and whether integrating SWE with bioimpedance spectroscopy (BIS), range of motion (ROM), and patient-reported outcomes (PROs) improves differentiation of MPS subgroups. Seventy-one adults completed upper-trapezius SWE, BIS, ROM assessments, and PRO measures. Clinically, 18 were classified as active MPS, 36 as latent, and 17 as normal. Shear wave speed measurements were modeled to estimate longitudinal (uL), transverse (uT), and anisotropy (uE) components. Reliability was examined using intraclass correlation coefficients. Unsupervised clustering and partial-correlation network analysis were applied to biomechanical and clinical variables. uT showed the strongest associations with BIS frequency parameters and ROM measures, indicating sensitivity to fascial composition, and mobility. Multimodal clustering incorporating uT with Fc or ROM identified subgroups with distinct tissue-level and functional characteristics. Network analysis demonstrated a progression in connectivity patterns, shifting from localized mechanical relationships to broader symptom-level coupling involving pain interference, sleep disturbance, emotional distress, and physical function. These findings indicate that SWE-derived stiffness parameters provide reliable, direction-specific quantification of trapezius mechanical properties. Combining SWE with impedance and mobility measures yields physiologically coherent MPS phenotypes that differ in both biomechanical features and clinical network structure, supporting more objective framework for characterizing MPS.

## I. INTRODUCTION

Myofascial pain syndrome (MPS) is a prevalent musculoskeletal condition characterized by myofascial trigger points (MTrPs), defined as hyperirritable nodules within taut bands of skeletal muscle that elicit referred pain, local tenderness, and motor dysfunction [1]. Clinically, MPS can present as active, latent, or normal states. Active MTrPs reproduce a patient’s characteristic pain upon palpation, latent MTrPs are tender but do not produce spontaneous pain, and normal muscle shows no taut bands or trigger point sensitivity [2]. Individuals with MPS frequently exhibit increased muscle stiffness, reduced range of motion (ROM), and functional limitation that contribute to substantial disability and healthcare burden [3], [4]. Despite its clinical significance, diagnosis remains largely subjective, relying on manual palpation and patient-reported symptoms. The absence of reliable, objective biomarkers limits the ability to classify patients, tailor interventions, and monitor treatment response [5].

Shear wave elastography (SWE) is emerging as a promising modality for quantitative assessment of muscle tissue properties[6], [7]. By quantifying shear wave propagation velocities, SWE enables non-invasive assessment of muscle and fascia stiffness and can provide insight into tissue anisotropy [8], [9]. Prior studies have reported increased stiffness in muscles harboring active trigger points or in regions of chronic myofascial pain [10], [11]. However, the clinical utility of SWE remains limited by persistent challenges in reproducibility and the difficulty of interpreting shear wave propagation through heterogeneous, anisotropic medium like muscle tissue. Most existing studies rely on single-angle or single-parameter measurements and rarely evaluate stiffness across different probe orientations or incorporate complementary physiological and functional metrics. In addition, variability in acquisition protocols and inter-operator reliability continues to restrict broader clinical adoption [12], [13]. At the same time, individuals with MPS exhibit substantial heterogeneity in both clinical presentation and tissue-level characteristics, indicating that single-parameter SWE alone cannot fully capture the complexity of the condition. These limitations collectively highlight the need for multi-directional SWE assessment integrated with complementary biomechanical and clinical measures to reveal meaningful physiological differences.

Complementary assessments such as bioimpedance spectroscopy (BIS), ROM as well as clinical assessments may help contextualize SWE findings by capturing structural and functional manifestations of MPS. BIS provides indirect markers extra- and intracellular fluid [16], features implicated in fascial densification, pain sensitization and local neurogenic inflammation. ROM, which is frequently restricted in MPS and improves with targeted interventions, captures the functional consequences of tissue stiffness and pain-related motor inhibition [17]. While each measure has limitations when used in isolation, integrating them with SWE may offer a more complete biomechanical characterization of myofascial dysfunction. Additionally, since these measures often co-occurs with distinct patterns of pain, emotional distress, and functional limitation, integrating them may reveal biomechanical profiles that align with clinically meaningful symptom patterns.

Considering these limitations, the primary objective of this study was to determine whether stiffness parameters obtained from SWE across multiple probe orientations can characterize clinically meaningful variability in MPS. Specifically, we examined whether SWE alone is sufficient to differentiate tissue properties across individuals with active, latent, and no myofascial pain, and whether combining SWE with BIS and ROM improves biomechanical and clinical subgrouping. Our contributions are twofold: (1) evaluating whether rotational SWE can provide reliable and physiologically interpretable estimates of upper-trapezius anisotropy, and (2) examining whether integrating SWE with impedance and mobility measures can reveal distinct biomechanical profiles that correspond to meaningful patterns of clinical symptom organization. Together, these goals seek to establish the potential role of multimodal ultrasound-based assessment in advancing objective, mechanism-informed classification of myofascial pain.

## II. METHODS

This study followed a multimodal analytical framework to integrate biomechanical, compositional, functional, and clinical information into a unified assessment pipeline. Data acquisition included clinical evaluation. (Section B.1), self-reported questionnaires (Section B.2), musculoskeletal and sensory evaluation (Section B.3), SWE with 360° rotational sweeps (Section B.4), ROM testing (Section B.5) and BIS (Section B.6). These multimodal data streams were then processed to extract rotational stiffness parameters from SWE (Section C.1). Reliability analyses were conducted to evaluate the stability of SWE-derived parameters across repeated angular sweeps (Section C.1). The processed feature sets were subsequently used for unsupervised clustering to identify biomechanical subgroups based on SWE alone and in combination with BIS or ROM (Section C.2). Finally, partial-correlation network modeling characterized interrelationships among clinical, sensory, and functional variables within each subgroup. This workflow links quantitative tissue properties with multidimensional clinical profiles in MPS.

### A. Study Design and Participants

This cross-sectional study was conducted at George Mason University as part of a broader research effort to identify objective biomechanical markers related to myofascial pain (registered on ClinicalTrials.gov: NCT06060925). Adults aged 18 years and older were invited to participate, including individuals reporting chronic neck and shoulder pain, as well as individuals without current symptoms. Recruitment was carried out through community outreach and regional private healthcare practices across Northern Virginia, Washington, D.C., and Baltimore.

Exclusion criteria included a diagnosis of fibromyalgia, chronic fatigue syndrome, chronic Lyme disease, cervical radiculopathy, neuropathy, or neuritis. Additional exclusions were prior surgery involving the head, neck, or shoulder girdle; medication changes in the past six months; or a history of cervical or shoulder fractures. All study procedures were approved by the university’s Institutional Review Board, and written informed consent was obtained prior to participation.

### B. Outcome Measures

#### 1. Standardized Clinical Evaluation

Each participant underwent a structured musculoskeletal evaluation conducted by two board-certified physicians and two licensed physical therapists. Based on clinical findings, participants were categorized into three groups:

- Active: characterized by spontaneous, ongoing pain in the affected region, often accompanied by referred pain, and increased sensitivity.
- Latent: asymptomatic at rest but containing palpable trigger points that elicit tenderness or referred pain only when stimulated.
- Normal: asymptomatic, with no palpable trigger points and no clinical signs of myofascial dysfunction.

While detailed trigger-point mapping was performed, the current analysis focused on quantifiable outcomes.

#### 2. Self-Reported Health and Pain Measures

Validated questionnaires based on the NIH HEAL Common Data Elements were used to quantify pain, function, emotional distress, and sleep quality: Pain, Enjoyment, and General Activity (PEG) [18]; Pain Catastrophizing Scale (PCS) [19]; PROMIS Physical Function [20]; PROMIS Sleep Disturbance [21]; Generalized Anxiety Disorder-2 (GAD-2) [22]; Patient Health Questionnaire-2 (PHQ-2) for depressive symptoms [23].

#### 3. Musculoskeletal and Sensory Function

Hypermobility was assessed with a modified Beighton– Brighton scale (19-point composite) incorporating standard Beighton criteria plus additional Brighton indicators [24]. Pressure Pain Threshold (PPT) was measured at the medial upper trapezius using a digital algometer (Commander Algometer, JTech Medical). The minimum pressure eliciting discomfort was recorded bilaterally. Quantitative sensory testing (QST) was performed using temporal summation as an indication of central sensitization. It was evaluated using 16 mechanical pinprick stimuli (256 mN, 1 Hz) bilaterally to the upper trapezius (PinPrick Stimulator Set, MRC Systems). Participants rated pain intensity after each stimulus on a 0–10 numeric rating scale [25].

#### 4. Ultrasound Imaging and Shear Wave Elastography

SWE was used to quantify the rotational stiffness of the upper trapezius muscle. Participants lay prone on an examination table with arms resting comfortably at their sides to minimize motion artifacts.

Bilateral SWE acquisitions were performed using a robotic arm (Universal Robots UR series) programmed to rotate the ultrasound probe (SL 10-2 linear array, SuperSonic Imagine, Aix-en-Provence, France) in 10° increments across a full 360° sweep around the muscle. The sweep began from the cranial orientation and proceeded clockwise, maintaining minimal probe pressure and consistent contact. For each acquisition, a fixed 5-cm region of interest (ROI) was centered over the muscle belly to ensure anatomical consistency across participants (Figure 1).

**Figure 1:**
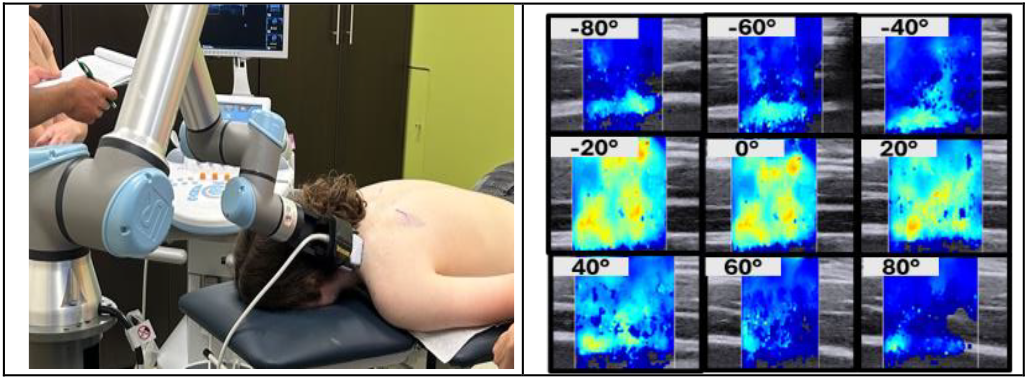
Example shear wave elastography image.

At each angular position, shear wave images were recorded once the elastogram stabilized. Images were displayed in meters per second (0–4.1 m/s) with uniform acquisition parameters (frequency: 2–10 MHz; dynamic range: 60 dB; persistence: medium; frame rate: 12 Hz). All images were visually inspected for artifacts (e.g., dropout, shadowing) and excluded if unstable.

For each participant, two full sweeps (0–180° and 180– 360°) were collected per side. Mean shear-wave speed values from each 10° increment were used to generate an angular stiffness profile. Profiles were normalized so that the peak speed aligned at 90°, corresponding to the fiber direction.

Angular SWE profiles and rotational stiffness modeling were based on established anisotropic elastic modeling in skeletal muscle [26]:

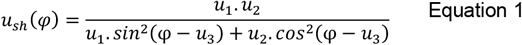

where *u*_*1*_ and *u*_*2*_ represent stiffness along and across fibers, and *u*_*3*_ denotes fiber orientation. From this model, three stiffness parameters were derived for each side:

- uL (longitudinal stiffness): stiffness along the fiber direction.
- uT (transverse stiffness): stiffness across the fiber direction.
- uE (anisotropy index): (uL – uT) / uL, expressing rotational contrast.

An example of a rotational stiffness profile and fitted model is shown in Figure 2, demonstrating clear angular dependence with a peak near the fiber direction.

**Figure 2.**
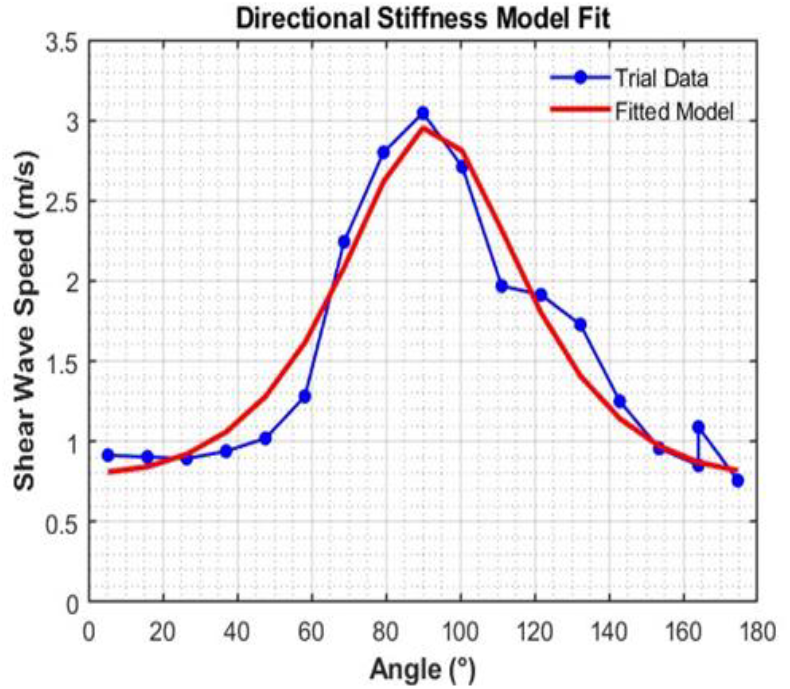
Shear wave speed profiles (blue) and fitted model (red).

#### 5. Range of Motion (ROM)

Shoulder and cervical joint ROM were assessed using a markerless 3D motion-capture system (Kinotek Inc., Portland, ME) with a Microsoft Azure Kinect depth-sensing camera. The system employs LiDAR and machine-learning algorithms to track anatomical landmarks without physical markers.

Participants, wearing fitted light-colored clothing, stood at a standardized distance from the sensor and followed on-screen prompts. Each 30-second trial captured the following movements: cervical side-bending (left/right); shoulder abduction (left/right); shoulder internal and external rotation (90° abduction, 90° elbow flexion). The maximum angle achieved for each motion per side was extracted and averaged across trials. Bilateral means were computed (e.g., *ROM_Shoulder_Flexion_Avg*) to reduce asymmetry and serve as functional input features for multimodal clustering.

#### 6. Bioimpedance Spectroscopy

Local tissue conductivity was evaluated using a single-channel tetrapolar BIS unit (SFB7, ImpediMed, QLD, Australia). Four adhesive ring electrodes were applied along a line connecting the acromion and C7 vertebra, centered on the mid-portion of the upper trapezius.

Each measurement performed a 256-frequency sweep (3 kHz – 1000 kHz) to generate impedance spectra, which were modeled using the Cole–Cole equation [27]. The following BIS-derived indices were extracted:

- BIS_Rlow: resistance at low frequency, reflecting extracellular fluid.
- BIS_Rhigh: resistance at high frequency, representing total tissue conductivity.
- BIS_fc (characteristic frequency): frequency of maximum reactance, inversely related to tissue hydration and cell permeability.
- BIS_Phi (phase angle): phase at fc, indicating cell integrity and dielectric behavior.

Bilateral BIS features were averaged (*Fc_avg, Phi_avg, Rlow_avg, Rhigh_avg*) for multimodal integration with SWE and ROM measures.

### C. Data Analysis

#### 1. Extraction and Reliability of SWE Parameters

Rotational stiffness profiles from each SWE scan were analyzed using the nonlinear cosine-based model described in Equation 1. Three parameters were derived for each side: longitudinal stiffness (*uL*), transverse stiffness (*uT*), and anisotropy index (*uE*). Model fit was evaluated separately for each participant by computing an individual coefficient of determination (R^2^), which reflects how well the angular stiffness measurements were captured by the rotational model for that subject. Because a separate model is estimated for each participant, R^2^ values were used only as descriptive indicators of individual fit quality and were summarized using the mean and standard deviation.

To assess measurement reproducibility across the two angular sweeps per side, single-measurement intra-class correlation coefficients (ICC) were computed. Reliability was interpreted according to established thresholds: poor (<0.5), moderate (0.5–0.75), good (0.75–0.90), and excellent (>0.90). Parameters showing good-to-excellent reliability (ICC ≥ 0.70) were retained for further analyses [28].

To complement ICC-based reliability assessment, Bland–Altman agreement analyses were also performed for each SWE-derived parameter [29]. For each subject and side, the mean difference (bias) and 95% limits of agreement (LoA = bias ± 1.96 × SD of the differences) between the two angular sweeps were calculated. Bland–Altman plots of the difference versus the average of the two sweeps were visually inspected to evaluate systematic bias and potential heteroscedasticity. This analysis provided an absolute measure of agreement to confirm the consistency of stiffness estimates across repeated rotational measurements.

#### 2. Clustering Analysis

##### Framework and Validation

Unsupervised clustering was used to identify biomechanical and physiological subgroups of participants. Analyses were conducted in two stages: (1) SWE-based clustering using stiffness parameters alone; and (2) multimodal clustering integrating SWE features with complementary measures (BIS and ROM).

All features were standardized before [30]. The optimal number of clusters (k) was determined using silhouette coefficients and visual inspection of cluster cohesion and separation[31]. Between-cluster differences were tested using Kruskal–Wallis tests, which yield an H statistic that follows an approximate χ^2^ distribution under the null hypothesis. These tests were used to assess whether clusters exhibited meaningful separation along the stiffness, BIS, or ROM dimensions[32]. To assess external validity, PEG scores were compared across clusters as an indicator of pain interference. Imaging-derived clusters were also compared with clinician-defined categories (Active, Latent, Normal) to examine alignment with clinical phenotypes.

##### SWE-Based Clustering

In the first stage, clustering was performed using SWE-derived stiffness parameters to determine whether mechanical features alone could distinguish tissue-level phenotypes. Longitudinal stiffness values (*uL_L, uL_R*) were extracted from the rotational SWE model for each participant. Because *uL* reflects stiffness along the muscle-fiber axis, the physiologically dominant direction, it was used as the primary input variable for this analysis.

##### Feature Correlation and Selection for Multimodal Integration

Before multimodal clustering, pairwise Spearman correlations were computed to identify physiologically meaningful associations between SWE parameters (uL, uT, uE) and complementary measures, including BIS-derived indices, fascial morphology descriptors, and ROM metrics. Variables showing the strongest and most interpretable associations with SWE features were selected as complementary features. This process guided the construction of SWE–complementary feature pairs used in multimodal clustering.

##### Multimodal Clustering

In the second stage, SWE parameters were combined with BIS and ROM measures to evaluate whether integrating mechanical, compositional, and functional features improved subgroup differentiation. Each feature pair was analyzed using the same PAM algorithm and validation framework described above[30]. For comparability with clinical groupings, a fixed three-cluster (k = 3) configuration was used for all multimodal models.

Model evaluation and selection for downstream network analysis were based on:

- Silhouette coefficient, reflecting cluster cohesion and separation [31].
- Cluster balance, requiring ≥5 participants per subgroup for statistical stability.
- Feature significance, based on Kruskal–Wallis tests for both stiffness and complementary variables. Just as a screening metric, we kept variables where p<0.05 [32].
- Anatomical consistency, favoring bilateral-averaged features (e.g., uT_avg, Fc_avg, ROM_avg) to minimize side bias.

Models satisfying all four criteria were retained for network characterization.

##### Network Analysis

##### Network Construction

To examine clinical organization within each biomechanical subgroup, network analyses were performed using regularized partial correlation networks estimated from clinical, sensory, and physical exam variables. Partial correlations quantify the direct conditional association between two variables after controlling for all remaining variables in the network, thereby isolating unique pairwise relationships. This approach captured interdependencies between pain, emotional distress, sensory sensitivity, and physical function. Each network included the following variables:

- Self-reported measures: PEG, PCS, PHQ-2, GAD-2, PROMIS Sleep Disturbance, and PROMIS Physical Function.
- Musculoskeletal and sensory measures: Pressure Pain Threshold (PPT), Quantitative Sensory Testing (QST), and Beighton hypermobility scores.

In Gaussian graphical models, Partial correlations are derived from the precision matrix (Θ), defined as the inverse of the covariance matrix (Σ) [33]. The partial correlation between variables i and j was computed as:

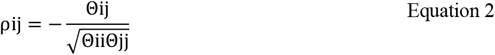

Where Θ_??_ denotes the (i,j) entry of the precision matrix. The negative sign ensures correct directionality of the resulting partial correlation, as required by Gaussian graphical theory [33], [34]. In this study, partial correlations and precision matrices were estimated directly using regularized Gaussian graphical modeling, as described below.

##### Graphical LASSO Regularization

The sparse precision matrix was estimated using the graphical least absolute shrinkage and selection operator (GLASSO), this method estimates Θ by solving the penalized maximum-likelihood optimization problem:

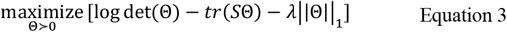

where:

- Θ is the precision matrix
- S is the empirical covariance matrix
- ∥Θ∥_1_ denotes the element-wise l1-norm (sum of absolute values of all entries)
- and log det (Θ)™tr(SΘ) represents the Gaussian log-likelihood.
- λ is the regularization parameter controlling sparsity.

Graphical LASSO yields a sparse estimate of conditional dependencies by shrinking small partial correlations to zero, thereby improving interpretability and reducing spurious associations [35]. Network estimation was performed using EBIC-regularized Graphical LASSO, which selects the optimal λ based on the Extended Bayesian Information Criterion (EBIC), balancing goodness-of-fit with model parsimony[36], [37]. In the resulting networks, variables are represented as nodes and conditional associations as edges, with edge thickness corresponding to the strength of the partial correlation.

##### Centrality Analysis

For each cluster-specific network, four standard graph-theoretic centrality metrics were computed, including strength, expected influence, closeness, and betweenness. Strength reflects the sum of absolute edge weights connected to a node, whereas expected influence preserves the signed contribution of all connected edges. Closeness quantifies the inverse of the average shortest path length to all other nodes, and betweenness reflects the number of shortest paths that pass through a node. These metrics are widely used in weighted psychological and clinical networks [36], [38], [39], [40]

##### Network Stability and Visualization

Network stability and robustness were evaluated using non-parametric case-dropping bootstrap procedures[36]. In this approach, networks were repeatedly re-estimated after randomly removing increasing proportions of participants (from 10% to 70%). Two stability metrics were examined: (1) edge-weight stability, quantified as the correlation between original and bootstrapped edge weights at each level of case removal; and (2) strength centrality stability, quantified as the correlation between original and re-estimated node strength values across bootstrap samples. Stability was assessed graphically using bootstrap correlation curves, which display the average correlation between original and re-estimated parameters as a function of the proportion of cases removed. Higher and more slowly decaying correlation curves indicate greater network robustness.

## III. RESULTS

### A. Participant Characteristics

A total of 96 subjects were enrolled in this study. After excluding participants with incomplete SWE, BIS, ROM or questionnaire data, 71 individuals had complete records and were included in the final analysis. The sample had a median age of 32 years (IQR = 14). Females comprised 58.6% of the sample and males 41.4%. Participants self-identified as Asian (35.7%), White (51.4%), and Black or African American (8.6%). Based on clinical examination, participants were categorized into Active MPS (n = 18), Latent MPS (n = 36), and Normal controls (n = 17). Descriptive statistics for all clinical and self-reported measures are summarized in Table 2.

**Table 1.**
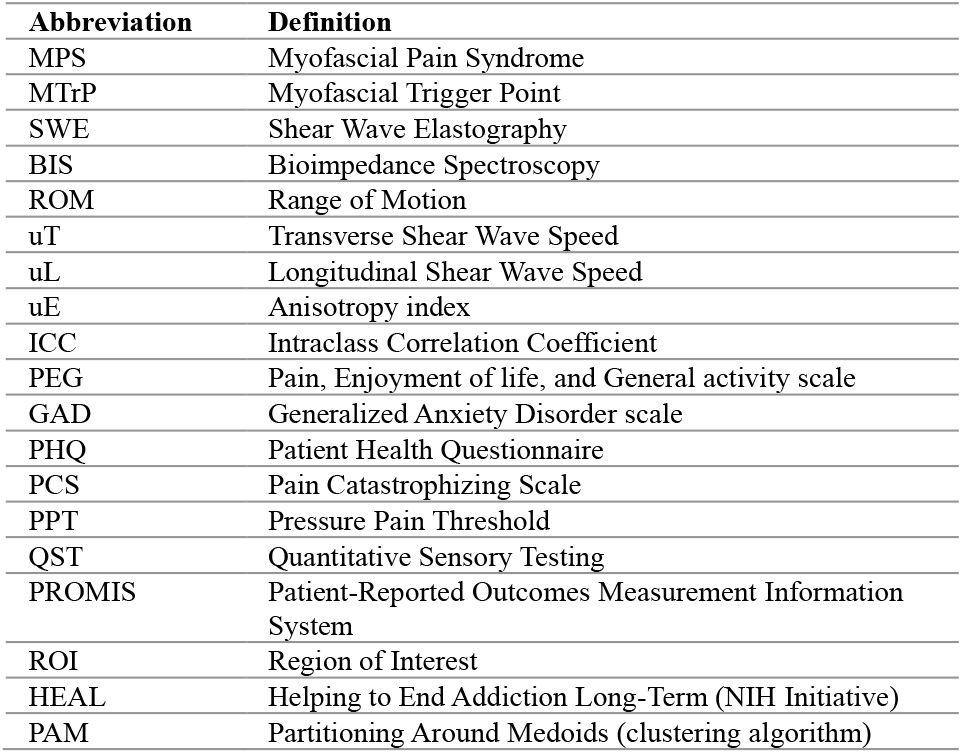
List of abbreviations.

| Abbreviation | Definition |
| --- | --- |
| MPS | Myofascial Pain Syndrome |
| MTrP | Myofascial Trigger Point |
| SWE | Shear Wave Elastography |
| BIS | Bioimpedance Spectroscopy |
| ROM | Range of Motion |
| uT | Transverse Shear Wave Speed |
| uL | Longitudinal Shear Wave Speed |
| uE | Anisotropy index |
| ICC | Intraclass Correlation Coefficient |
| PEG | Pain, Enjoyment of life, and General activity scale |
| GAD | Generalized Anxiety Disorder scale |
| PHQ | Patient Health Questionnaire |
| PCS | Pain Catastrophizing Scale |
| PPT | Pressure Pain Threshold |
| QST | Quantitative Sensory Testing |
| PROMIS | Patient-Reported Outcomes Measurement Information System |
| ROI | Region of Interest |
| HEAL | Helping to End Addiction Long-Term (NIH Initiative) |
| PAM | Partitioning Around Medoids (clustering algorithm) |

**Table 2.**
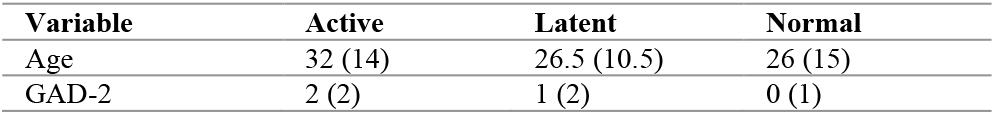
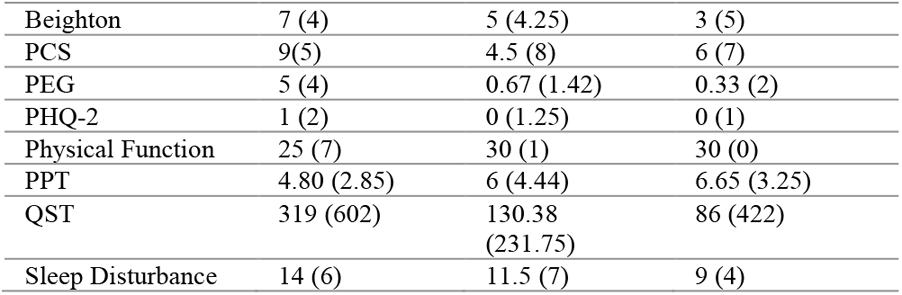
Sample characteristics by MPS group. Values are presented as median (IQR).

### B. SWE Modeling and Reliability Assessment

The nonlinear rotational stiffness model provided a strong fit to the angular SWE profiles across participants. The average model fit quality was high, with a median R^2^ of 0.83 (IQR=0.22), indicating good agreement between measured and fitted values. Group-level stiffness parameters were summarized using median (IQR): longitudinal stiffness uL = 2.37 m/s (1.12), transverse stiffness uT = 0.94 m/s (0.58), and anisotropy index uE = 0.59 (0.28).

Reliability analyses demonstrated moderate to excellent agreement across repeated sweeps. Intra-class correlation coefficients (ICCs) for uL, uT, and uE are summarized in Table 3. Overall, uL and uT exhibited high reproducibility across trials, while uE showed lower yet acceptable stability.

**Table 3.** SWE model fit, group-level stiffness parameters, and reliability.

| Parameter | Mean ± SD | ICC (Left, 95% CI) | ICC (Right, 95% CI) |
| --- | --- | --- | --- |
| $uL$ | $2.41 \pm 0.81$ | 0.76 (0.66–0.83) | 0.79 (0.69–0.85) |
| $uT$ | $1.06 \pm 0.60$ | 0.89 (0.83–0.92) | 0.89 (0.84–0.93) |
| $uE$ | $0.55 \pm 0.19$ | 0.64 (0.50–0.75) | 0.61 (0.46–0.72) |

**Table 4.** Centrality measures for clusters obtained from uL_L + uL_R. BC = Betweenness Centrality; CC = Closeness Centrality; EI = Expected Influence.

| Node | BC | CC | Strength | EI |
| --- | --- | --- | --- | --- |
| **Cluster 1** |  |  |  |  |
| PHQ 2 Total Score | 1 | 0.014 | 0.782 | 0.595 |
| GAD 2 Total Score | 8 | 0.017 | 0.968 | 0.216 |
| PCS Total Score | 3 | 0.015 | 0.770 | 0.082 |
| PEG Score | 0 | 0.013 | 0.871 | 0.033 |
| PROMIS Physical Function | 15 | 0.017 | 1.178 | -1.178 |
| PROMIS Sleep Disturbance | 0 | 0.015 | 0.651 | 0.354 |
| QST Sumsquare | 1 | 0.012 | 0.597 | 0.276 |
| Combined PPT AVG | 1 | 0.013 | 0.795 | -0.348 |
| Beighton total | 0 | 0.010 | 0.332 | -0.084 |
| **Cluster 2** |  |  |  |  |
| PHQ 2 Total Score | 0 | 0.55 | 0.550 | 0.550 |
| GAD 2 Total Score | 0 | 0.55 | 0.550 | 0.550 |
| PCS Total Score | 0 | NA | 0.000 | 0.000 |
| PEG Score | 0 | NA | 0.402 | -0.402 |
| PROMIS Physical Function | 0 | NA | 0.402 | -0.402 |
| PROMIS Sleep Disturbance | 0 | NA | 0.000 | 0.000 |
| QST Sumsquare | 0 | NA | 0.451 | -0.451 |
| Combined PPT AVG | 0 | NA | 0.451 | -0.451 |
| Beighton total | 0 | NA | 0.000 | 0.000 |

To further assess agreement between repeated trials, Bland–Altman analyses were performed for uL and uT on both sides (Figure 3). The plots revealed that the mean bias between trials was negligible for both parameters (uL: +0.02 m/s [left], –0.07 m/s [right]; uT: +0.01 m/s [left], +0.03 m/s [right]). The differences were randomly distributed around zero without evidence of proportional bias, and over 95 % of the observations fell within the ±1.96 SD limits of agreement. These findings confirm that the rotational stiffness parameters derived from the SWE model are highly repeatable and show excellent intra-session reliability for both sides of the trapezius muscle.

**Figure 3.**
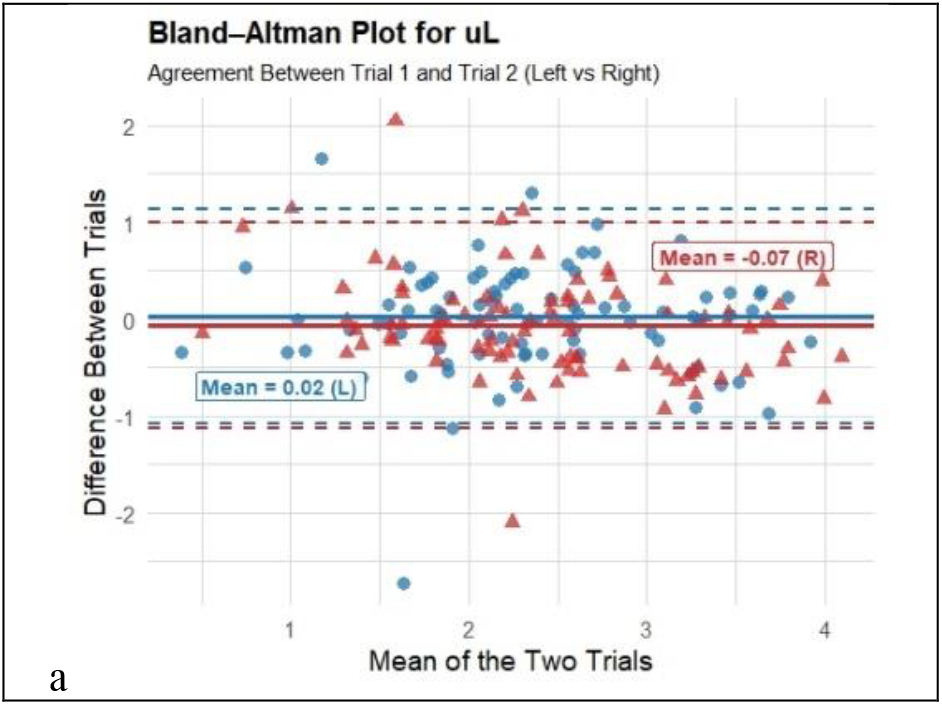
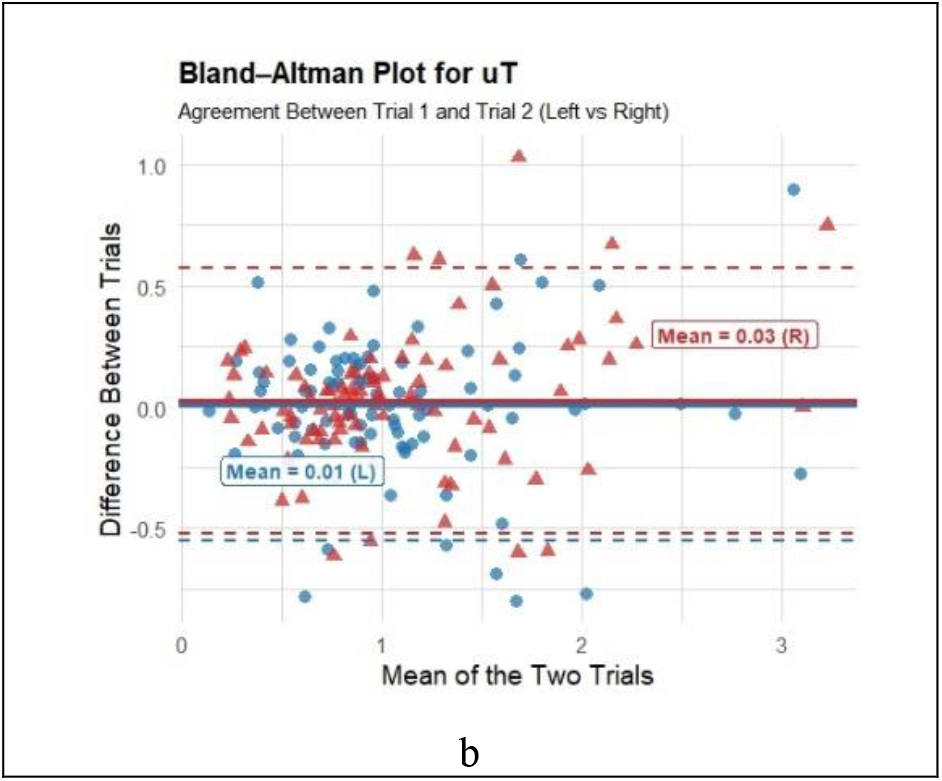
Bland–Altman plots evaluating repeatability for (a) longitudinal (uL) and (b) transverse (uT) shear speed.

### C. SWE-Based Grouping of Clinical Phenotypes

#### 1. Clustering and Validation

Cluster analysis was performed using standardized longitudinal stiffness values (uL_L and uL_R). Although uT demonstrated slightly higher reliability and lower variability, uL was chosen as the most meaningful feature for clustering because it directly reflects physiologically relevant stiffness along the muscle fiber direction.

A two-cluster model (k = 2) provided the clearest separation of participants based on bilateral longitudinal stiffness. Cluster 1 was characterized by consistently higher uL_L and uL_R values, whereas Cluster 2 exhibited lower values bilaterally. Cluster labels were compared with clinical categories (Active, Latent, Normal). Cluster 1 included 6 Active, 18 Latent, and 7 Normal participants, while Cluster 2 contained 13 Active, 19 Latent, and 8 Normal individuals. Scatterplots (Figure 4-a, and -b) confirmed distinct distributions, and Kruskal–Wallis tests indicated significant between-cluster differences for both uL_L and uL_R (p < 0.001), verifying that longitudinal stiffness reliably distinguished the groups.

**Figure 4.**
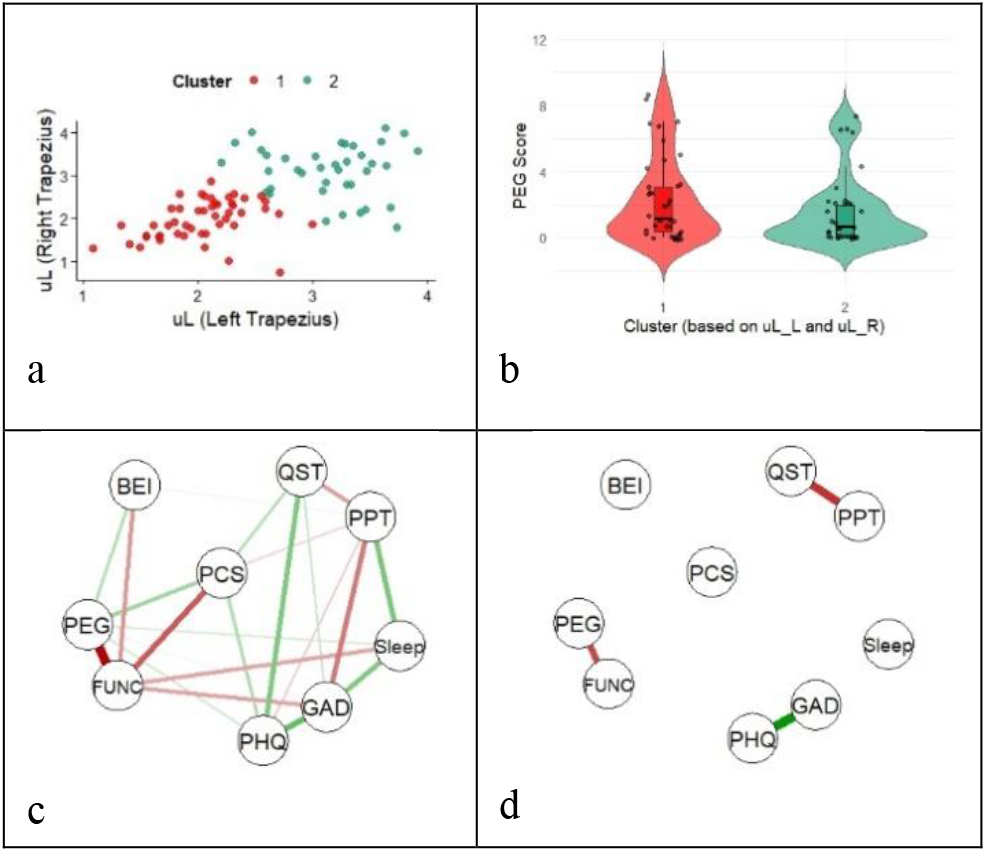
uL-based clustering with associated pain and network features: (a) bilateral uL scatterplot, (b) PEG score differences, (c) Cluster 1 network, and (d) Cluster 2 network.

Consistent with these findings, differences in pain interference further distinguished the two clusters. Violin-plot analysis (Figure 4-b) showed that Cluster 1, the high-stiffness group, had higher median PEG scores and greater variability, whereas Cluster 2 exhibited lower, more narrowly distributed scores.

#### 2. Clinical Distribution and Network Characterization

Partial-correlation network analyses were then conducted within each cluster to examine how pain, function, and psychosocial variables were organized (Figure 4-c, and -d).

Cluster 1 revealed a densely connected network that organized into two functional domains. The first domain grouped PHQ-2, GAD-2, Sleep, QST, and PPT, forming an emotional–sensory network. The second domain linked PCS, PEG, Physical Function, and Beighton, outlining a psychosocial–functional network. Centrality analysis showed that Physical Function had the highest betweenness (15) and closeness (1.178), indicating a key bridging role between emotional and functional subsystems. The stability plot indicated consistent edge reliability down to approximately 30% resampling, supporting the robustness of this organizational structure.

In contrast, Cluster 2 displayed a sparsely connected network with only three small links. PHQ-2 and GAD-2 formed a tightly coupled but isolated affective pair, indicating strong covariance between depression and anxiety without cross-links to pain or function. PEG connected solely with Physical Function, and PPT was related only to QST, producing two separate micro-modules representing psychophysical and functional domains. Centrality measures confirmed minimal interdependence across the network: betweenness and strength were near zero for all nodes, while PHQ-2 and GAD-2 had modest closeness (0.55), reflecting their localized association. Edge-stability plots showed moderate robustness, with key edges persisting until roughly 50% of the sample was resampled.

### D. Multimodal Grouping Integrating SWE and Complementary Measures

#### 1. Correlation Analysis

Correlation analyses were performed to investigate the relationships between SWE parameters (uL, uT) and complementary measures including bioimpedance spectroscopy (BIS) and range of motion (ROM) features. Across participants, longitudinal stiffness (uL) and its averaged value (uL_avg) did not show statistically detectable correlations with BIS or ROM parameters. In contrast, transverse stiffness (uT) demonstrated small-to-moderate correlations.

Both the left- and right-sided transverse measures were correlated with BIS frequency-related parameters (e.g., Fc_L, Fc_avg, rhigh_R), indicating that greater tissue stiffness corresponded to lower frequency conduction. Conversely, uT was positively correlated with BIS phase angle, with stronger effects for right-sided and averaged measures (uT_R: ρ = 0.330; uT_avg: ρ = 0.303). Furthermore, uT_avg was positively correlated with shoulder flexion ROM (uT_avg: ρ = 0.327). Therefore, uT, BIS, and shoulder flexion ROM were retained for subsequent multimodal clustering analyses. The statistically significant relationships are summarized in Table 5.

**Table 5.** Significant correlations (p < 0.05) found between SWE and BIS and ROM measures.

| SWE Parameter | Outcome Measure | Spearman's $\rho$ | p-value |
| --- | --- | --- | --- |
| uT_L | Fc_L | -0.297 | 0.0058** |
| uT_R | rhigh_R | -0.253 | 0.0182* |
| uT_R | Phi_R | +0.330 | 0.0019** |
| uT_avg | Fc_avg | -0.316 | 0.0029** |
| uT_avg | Phi_avg | +0.303 | 0.0044** |
| uT_avg | ROM Shoulder Flexion Avg | +0.327 | 0.0029** |

#### 2. Multimodal Clustering and Validation

To assess whether combining SWE with complementary physiological measures enhances subgroup differentiation, multimodal clustering analyses were performed using transverse stiffness (uT) integrated with BIS and shoulder-flexion ROM parameters.

A fixed three-cluster (k = 3) solution was applied to maintain comparability with clinical categories (Active, Latent, Normal). Table 6 summarizes the clustering outcomes, with silhouette coefficients indicated moderate subgroup separation. Two averaged models were selected as the most reliable and physiologically interpretable:

- uT_avg + Fc_avg: yielded balanced clusters and strong between-cluster differentiation for both stiffness and BIS frequency (*χ^2^ ≈ 34*.*81; p < 0*.*001*).
- uT_avg + ROM_Shoulder_Flexion_Avg: showed robust separation across mechanical and functional dimensions despite a lower silhouette coefficient (0.38).

**Table 6.** Multimodal clustering results combining transverse speed (uT) with complementary measures.

| Clustering Features | Silhouette Score | $\chi^2$ (Complement) | Clinical Distribution (Active / Latent / Normal) |
| --- | --- | --- | --- |
| $uT\_L + \Phi_{hi\_L}$ | 0.61 | 5.80 | C1: 1/1/0;<br>C2: 4/9/6;<br>C3: 13/26/11 |
| $uT\_L + Fc\_L$ | 0.42 | 27.87 | C1: 4/8/5;<br>C2: 10/24/9;<br>C3: 4/4/3 |
| $uT\_L + r_{high\_L}$ | 0.51 | 3.62 | C1: 14/26/11;<br>C2: 4/9/6;<br>C3: 0/1/0 |
| $uT\_R + \Phi_{hi\_R}$ | 0.59 | 6.95 | C1: 1/1/0;<br>C2: 4/11/2;<br>C3: 13/24/15 |
| $uT\_R + r_{high\_R}$ | 0.56 | 4.69 | C1: 14/24/15;<br>C2: 4/11/2;<br>C3: 0/1/0 |
| $uT\_avg + \Phi_{hi\_avg}$ | 0.61 | 6.01 | C1: 1/1/0;<br>C2: 3/11/2;<br>C3: 14/24/15 |
| $uT\_avg + Fc\_avg$ | 0.38 | 34.81 | C1: 7/16/9;<br>C2: 1/5/2;<br>C3: 10/15/6 |
| $uT\_avg + ROM\_Shoulder\_Flexion\_Avg$ | 0.38 | 43.78 | C1: 6/21/8;<br>C2: 3/11/1;<br>C3: 9/4/8 |

**Table 7.** Centrality measures for Clusters obtained from uT_avg + Fc_avg. BC = Betweenness Centrality; CC = Closeness Centrality; EI = Expected Influence.

| Node | BC | CC | Strength | EC |
| --- | --- | --- | --- | --- |
| **Cluster 1** |  |  |  |  |
| PHQ 2 Total Score | 0 | 0.159 | 0.609 | 0.609 |
| GAD 2 Total Score | 1 | 0.214 | 0.940 | 0.940 |
| PCS Total Score | 0 | NA | 0.000 | 0.000 |
| PEG Score | 0 | NA | 0.559 | -0.559 |
| PROMIS Physical Function | 0 | NA | 0.559 | 0.559 |
| PROMIS Sleep Disturbance | 0 | 0.130 | 0.331 | 0.331 |
| QST Sumsquare | 0 | NA | 0.433 | -0.433 |
| Combined PPT_AVG | 0 | NA | 0.433 | -0.433 |
| Beighton total | 0 | NA | 0.000 | 0.000 |
| **Cluster 2** |  |  |  |  |
| PHQ 2 Total Score | 0 | NA | 0.000 | 0.000 |
| GAD 2 Total Score | 0 | NA | 0.000 | 0.000 |
| PCS Total Score | 2 | 0.122 | 0.889 | 0.889 |
| PEG Score | 2 | 0.122 | 1.124 | -0.189 |
| PROMIS Physical Function | 0 | 0.089 | 0.657 | -0.657 |
| PROMIS Sleep Disturbance | 0 | NA | 0.000 | 0.000 |
| QST Sumsquare | 0 | 0.077 | 0.422 | 0.422 |
| Combined PPT_AVG | 0 | NA | 0.000 | 0.000 |
| Beighton total | 0 | NA | 0.000 | 0.000 |
| **Cluster 3** |  |  |  |  |
| PHQ 2 Total Score | 0 | NA | 0.000 | 0.000 |
| GAD 2 Total Score | 0 | NA | 0.487 | -0.487 |
| PCS Total Score | 0 | NA | 0.000 | 0.000 |
| PEG Score | 0 | 0.068 | 0.413 | 0.413 |
| PROMIS Physical Function | 2 | 0.101 | 0.874 | -0.874 |
| PROMIS Sleep Disturbance | 2 | 0.101 | 0.775 | 0.775 |
| QST Sumsquare | 0 | NA | 0.000 | 0.000 |
| Combined PPT_AVG | 0 | NA | 0.487 | -0.487 |
| Beighton total | 0 | 0.072 | 0.512 | -0.512 |

**Table 8.** Centrality measures for clusters obtained from uT_avg + Shoulder_Flexion_Avg. BC = Betweenness Centrality; CC = Closeness Centrality; EI = Expected Influence.

| Node | BC | CC | Strength | EC |
| --- | --- | --- | --- | --- |
| **Cluster 1** |  |  |  |  |
| PHQ 2 Total Score | 0 | 0.039 | 0.419 | -0.419 |
| GAD 2 Total Score | 0 | 0.053 | 0.907 | 0.072 |
| PCS Total Score | 0 | NA | 0.368 | 0.368 |
| PEG Score | 1 | NA | 0.977 | -0.242 |
| PROMIS Physical Function | 0 | NA | 0.609 | -0.609 |
| PROMIS Sleep Disturbance | 4 | 0.053 | 1.216 | 0.394 |
| QST Sumsquare | 0 | 0.035 | 0.411 | -0.411 |
| Combined PPT_AVG | 7 | 0.062 | 1.530 | -0.900 |
| Beighton total | 0 | 0.037 | 0.379 | -0.379 |
| **Cluster 2** |  |  |  |  |
| PHQ 2 Total Score | 2 | 0.152 | 1.467 | 0.099 |
| GAD 2 Total Score | 0 | 0.085 | 0.387 | 0.387 |
| PCS Total Score | 2 | NA | 0.922 | 0.922 |
| PEG Score | 2 | NA | 1.077 | -0.086 |
| PROMIS Physical Function | 0 | NA | 0.582 | -0.582 |
| PROMIS Sleep Disturbance | 0 | NA | 0.000 | 0.000 |
| QST Sumsquare | 0 | NA | 0.426 | 0.426 |
| Combined PPT_AVG | 0 | 0.129 | 1.129 | -0.239 |
| Beighton total | 0 | 0.101 | 0.841 | 0.841 |
| **Cluster 3** |  |  |  |  |
| PHQ 2 Total Score | 0 | 0.168 | 0.679 | 0.679 |
| GAD 2 Total Score | 1 | 0.223 | 1.012 | 0.347 |
| PCS Total Score | 0 | NA | 0.435 | -0.435 |
| PEG Score | 0 | NA | 0.353 | -0.353 |
| PROMIS Physical Function | 1 | NA | 0.788 | -0.788 |
| PROMIS Sleep Disturbance | 0 | NA | 0.000 | 0.000 |
| QST Sumsquare | 0 | NA | 0.000 | 0.000 |
| Combined PPT AVG | 0 | 0.134 | 0.332 | -0.332 |
| Beighton total | 0 | NA | 0.000 | 0.000 |

Scatter plots (Figure 5-a) illustrate participant distribution across clusters. In the uT_avg + Fc_avg model, clusters separated clearly along stiffness and BIS axes, whereas in the uT_avg + ROM_Shoulder_Flexion_Avg model, separation occurred along stiffness and functional axes.

**Figure 5:**
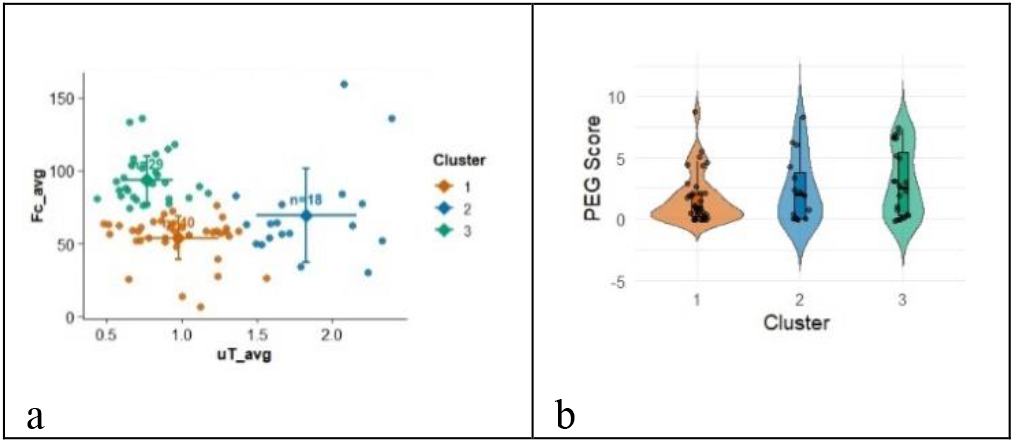
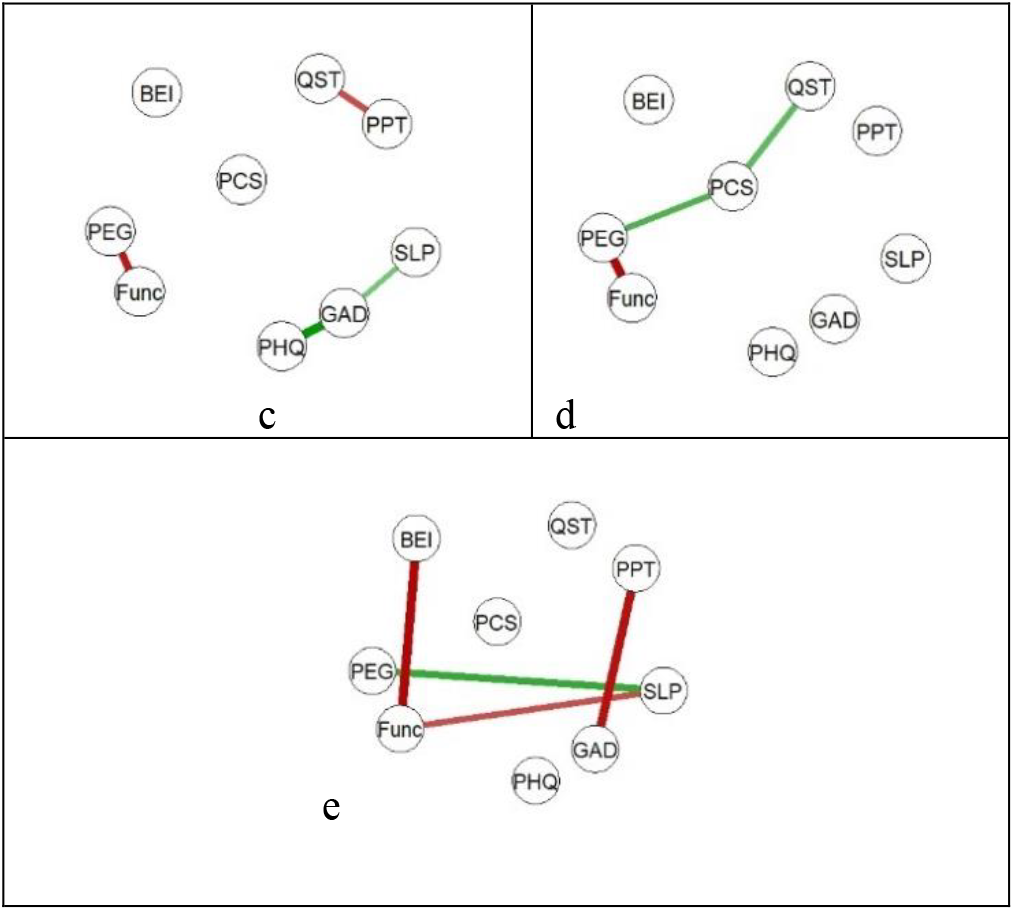
Clustering based on uT_avg and Fc_avg: (a) three data-driven clusters, (b) PEG score differences across clusters, and (c–e) partial-correlation networks for Clusters 1, 2, and 3.

The distribution of PEG (pain interference) scores across clusters derived from both multimodal models revealed consistent yet model-specific patterns of variation (Figure 5-b). In the uT_avg + Fc_avg model, Cluster 1 exhibited the lowest median PEG scores with narrow variability, Cluster 2 showed higher and more dispersed values, and Cluster 3 demonstrated the highest median PEG levels. This pattern indicates a graded increase in pain interference from Cluster 1 to Cluster 3, aligning with biomechanical differentiation along the stiffness–frequency axis. Conversely, in the uT_avg + ROM_Shoulder_Flexion_Avg model, Cluster 1 showed the highest median PEG scores and greatest variability, Cluster 2 had the lowest and most consistent values, and Cluster 3 intermediate levels. These findings suggest a graded decline in self-reported pain interference from Cluster 1 to Cluster 2, with Cluster 3 occupying a transitional position.

#### 3. Network Characterization of Multimodal Clusters

*(a)uT_avg + Fc_avg Model*

Partial-correlation network analyses (Figure 5-c, -d, and -e) revealed distinct interrelationships across the three biomechanical subgroups.

- Cluster 1: Sparse network with localized associations, PEG–Function formed a pain–function axis, while PHQ-2–GAD-2–Sleep formed an emotional–fatigue triad. PPT–QST captured sensory coupling. Most other nodes (e.g., PCS, Beighton) were isolated. Centrality values were low (Strength < 0.5), and edge stability declined below 40 % resampling.
- Cluster 2: Sparse network linking Function, PEG, PCS, and QST, reflecting sensory–psychosocial integration. Within this configuration, PCS and PEG exhibited the highest betweenness and strength, serving as bridges between sensory and functional domains. The network demonstrated stable inter-variable associations, with edge reliability persisting up to approximately 65% resampling.
- Cluster 3: Exhibited a moderately integrated network. PEG, Sleep, Function, and Beighton formed a pain interference–functional limitation cluster. Additionally, GAD-2 and PPT were linked through Sleep, indicating affective–sensorimotor coupling. Function and Sleep showed the highest betweenness (2) and closeness (∼0.10), acting as central hubs coordinating emotional and functional domains. Edge-stability results confirmed that most connections remained consistent up to approximately 65% resampling.

*(b)uT_avg + ROM_Shoulder_Flexion_Avg Model*

Networks derived from the uT_avg + ROM_Shoulder_Flexion_Avg model (Figure 6-c, -d, and -e) revealed progressive coordination among sensory, emotional, and functional domains.

**Figure 6:**
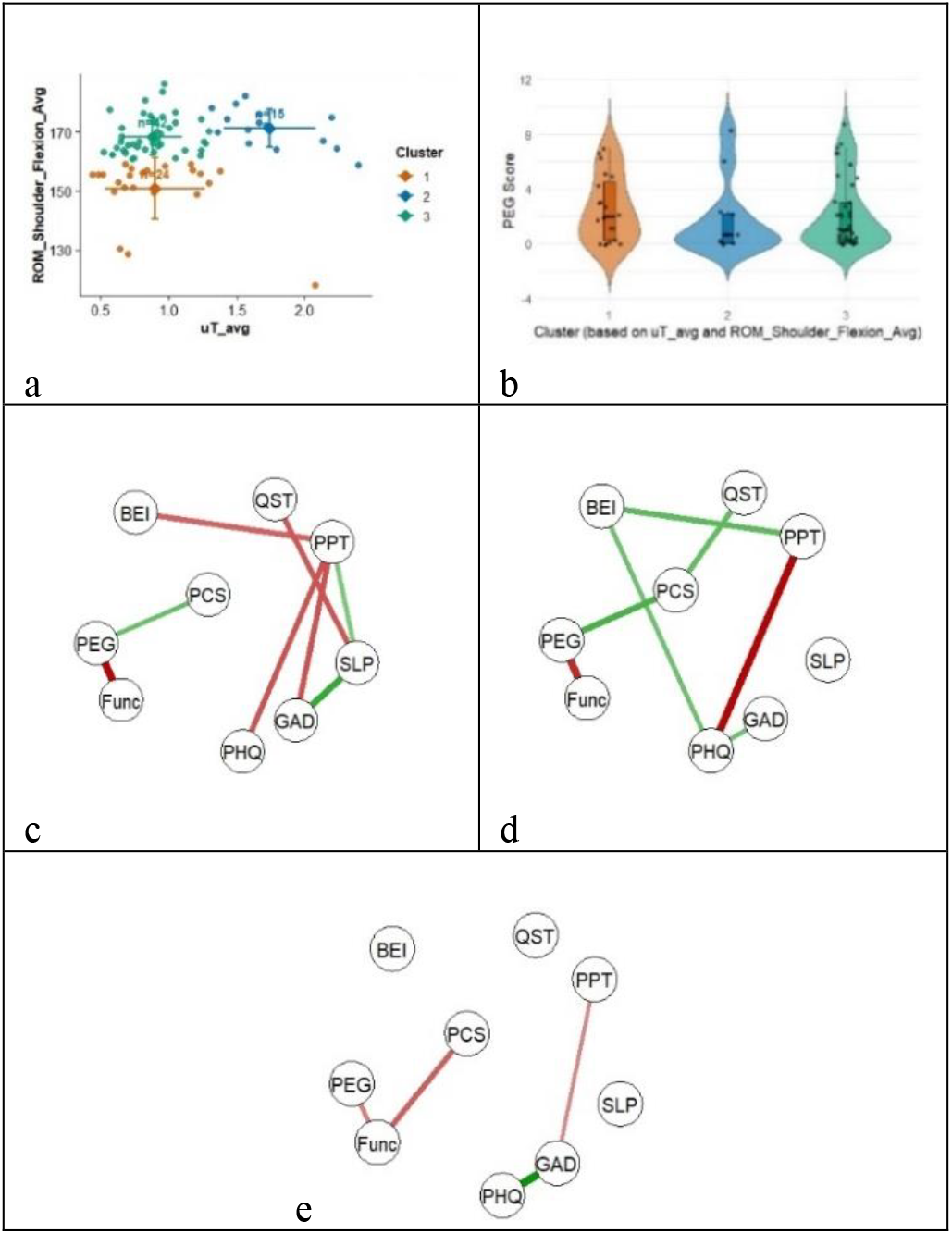
Clustering based on uT_avg and ROM_Shoulder_Flexion_Avg: (a) three data-driven clusters, (b) PEG score differences across clusters, and (c–e) partial-correlation networks for Clusters 1, 2, and 3.

- Cluster 1: Displayed a moderately integrated network centered on Sleep, which bridged emotional, sensory, and functional variables. A strong Function–PEG connection defined a clear pain–function axis, while the PPT–PHQ-2 link captured affective–sensory coupling. Sleep demonstrated the highest betweenness (4) and strength (1.22), serving as the key hub connecting psychological and physical dimensions. Edge-stability analysis indicated that most inter-variable associations remained consistent down to approximately 30% resampling.
- Cluster 2: Exhibited a cohesive configuration linking QST, PCS, PEG, and Function, forming a pain-catastrophizing module. Additional connections among PHQ-2, PPT, Beighton, and GAD-2 integrated emotional and sensory components into this broader framework. Centrality analysis revealed that PHQ-2, PPT, PEG, and PCS demonstrated the highest centrality values (Strength ≥ 0.9), indicating their pivotal roles as bridges between psychosocial and sensory–functional domains. Edge-stability results showed that correlations remained reliable up to approximately 40% resampling.
- Cluster 3: Presented a sparse network dominated by a small GAD-2–PHQ-2–PPT triad, accompanied by a secondary PEG–Function–PCS link. Within this structure, GAD-2 (Betweenness = 1; Strength = 1.01) and Function (Betweenness = 1; Strength = 0.79) emerged as primary hubs, facilitating limited interaction across emotional and functional domains. Edge-stability remained stable until approximately 30% resampling.

## IV. DISCUSSION

This study aimed to determine whether rotational SWE stiffness parameters, alone or in combination with BIS and ROM, can characterize clinically meaningful variability in individuals with myofascial pain syndrome. Our findings demonstrate that modeling anisotropic stiffness across 360° probe rotations yields reliable, direction-specific estimates of muscle and fascial mechanics. However, stiffness alone, particularly longitudinal stiffness, offered limited ability to explain the observed clinical heterogeneity. Integrating SWE with complementary measures such as BIS and ROM revealed more coherent biomechanical subgroups and clearer trends in symptom organization. These multimodal clusters exhibited distinct network patterns reflecting progressively stronger interactions among pain, emotional distress, sleep disturbance, and sensory sensitivity. Together, these findings show that MPS is best understood as a continuum of mechanical and clinical adaptation, and that combining quantitative tissue imaging with network-based analytics can support more precise, mechanism-informed phenotyping.

Following this overview, the Discussion is organized into six parts. Section 4.1 interprets the reliability and physiological meaning of rotational SWE metrics. Section 4.2 explains how integrating SWE with BIS and ROM improved biomechanical subgrouping. Section 4.3 examines the clinical networks within each subgroup and how symptom connectivity changes across mechanical profiles. Section 4.4 translates these biomechanical and network findings into clinical and pathophysiological implications, and Section 4.5 highlights future research directions, including longitudinal designs and precision rehabilitation.

### A. Reliability and Physiological Interpretation of Rotational SWE Metrics

The 3D rotational SWE protocol produced reproducible stiffness estimates for the upper trapezius, with longitudinal and transverse parameters showing good-to-excellent reliability (ICC = 0.76–0.89). Although the two 180° sweeps originate from the same continuous rotation rather than fully independent trials, the level of consistency still indicates that, under standardized acquisition, rotational SWE can stably characterize the expected anisotropic behavior of muscle tissue. The stability of the cosine-based model (with a median R^2^ of 0.83 (IQR = 0.22)) further supports its ability to align angular profiles with true fiber orientation instead of relying on single-angle measurements.

Physiologically, the model-derived parameters, uL (longitudinal stiffness), uT (transverse stiffness), and uE (anisotropy), represent distinct structural components of the muscle–fascia system. Consistent with prior elastography and MR studies of muscle anisotropy, uL reflects fiber-aligned stiffness, uT captures the mechanical properties of fascia, connective layers, and interstitial fluid, and uE quantifies rotational contrast [7], [41].

Among these, uT emerged as the most consistently associated measure across our analyses. It showed significant associations with BIS-derived characteristic frequency (Fc) and phase angle (Phi), as well as with shoulder-flexion ROM. Higher uT corresponded to lower Fc and higher Phi, patterns indicative of reduced tissue conductivity, and was also associated with reduced mobility. These results suggest that cross-fiber stiffness is sensitive to fascial composition and collagen organization, aligning with prior work showing that chronic myofascial pain is linked to thickened, less compliant fascia [42].

In contrast, uL and uE demonstrated minimal relationships with complementary measures, indicating that changes across fibers better reflect early tissue remodeling in myofascial pain, or may be confounded by tension in the muscle fibers. Similar observations have been reported in elastography studies of lumbar and plantar fascia, where elevated transverse stiffness is linked to movement restriction and pain intensity [43].

Overall, rotational SWE enhanced both measurement reliability and physiological interpretability by separating fiber-aligned stiffness (uL) from transverse stiffness (uT) governed by fascia and connective tissue [16].

### B. Multimodal Clustering and Biomechanical Subgrouping

Multimodal clustering integrating transverse stiffness with bioimpedance and shoulder range of motion revealed distinct biomechanical subgroups that reflect different patterns of mechanical and functional adaptation in myofascial pain. Across all combinations, the uT + Fc and uT + shoulder-flexion ROM models produced the clearest and most physiologically meaningful separation. Both showed significant between-cluster differences (χ^2^ = 34–54, p < 0.001), indicating that tissue stiffness, conductivity, and mobility change together in MPS. Averaging stiffness across sides improved cluster stability and minimized unilateral bias. In the uT + Fc model, three subgroups emerged along a gradient of fascial conductivity and mechanical stiffness. One subgroup showed moderate stiffness with low Fc, suggesting reduced fascial mobility and possible fluid accumulation. A second subgroup displayed high stiffness with moderate Fc, consistent with denser fascia but partially preserved conductivity, likely representing a transitional stage. A third subgroup demonstrated low stiffness and high Fc, reflecting more compliant and well-hydrated tissue. These patterns align with evidence that chronic myofascial pain involves thickened, less conductive fascia as collagen crosslinking and fluid retention increase [16], [42].

The uT + shoulder-flexion ROM model similarly identified subgroups capturing the interaction between stiffness and movement. Individuals with low stiffness but restricted ROM likely reflect early local mechanical tightness. Those with high stiffness and preserved ROM may rely on compensatory motor strategies that maintain movement despite elevated tension. A third subgroup with low-to-moderate stiffness and wide ROM likely represents more flexible tissue with minimal restriction. Together, these patterns show that transverse stiffness and ROM jointly capture both structural and functional aspects of myofascial changes.

Importantly, the multimodal clusters aligned more closely with the clinical network patterns than the SWE-only clusters. Subgroups defined by stiffness alone showed sparse networks, with weak associations between pain, function, and emotional symptoms. In contrast, the multimodal subgroups exhibited more interconnected clinical networks, with stronger coupling among pain interference, emotional distress, sleep disturbance, and sensory sensitivity, features consistent with early central sensitization. The moderate silhouette values (0.33–0.42) support that combining SWE with functional or compositional measures yields more coherent and physiologically grounded classifications than stiffness alone.

Overall, these findings suggest that myofascial pain reflects a continuum of biomechanical states rather than a binary stiff-versus-normal pattern. Transverse stiffness, fascial conductivity, and joint mobility together characterize how tissues progress from localized restriction to more globally integrated symptom patterns. This multimodal perspective may provide a useful basis for tailoring interventions, emphasizing mechanical therapies for stiffness-dominant profiles and combined physical–psychological approaches for more complex or sensitized presentations.

### C. Network-Level Patterns and Symptom Integration

The network findings illustrate how different biomechanical profiles correspond to distinct patterns of symptom organization in myofascial pain. Across the three clustering approaches, SWE-only using uL, and multimodal models integrating uT with Fc or ROM, each subgroup showed a characteristic pattern of clinical connectivity, reflecting different clinical presentations of mechanical and psychosocial involvement.

Clustering based on longitudinal stiffness produced two mechanical subgroups that differed mainly in fiber-aligned shear speed. Cluster 1, the high-stiffness group, exhibited a dense clinical network linking pain interference, emotional distress, sleep disturbance, and function, suggesting that increased stiffness may be associated with coupling across psychosocial and functional domains. Cluster 2, with lower stiffness, showed a sparse and weakly connected network, consistent with a more localized or early mechanical stage in which symptoms remain relatively independent. Thus, the uL model appears to represent an early, fiber-dominant mechanical stage where only some individuals show emerging multidomain symptom interaction.

Integrating transverse stiffness with fascial conductivity revealed three subgroups that aligned more consistently with clinical network patterns. The moderate-stiffness/low-Fc subgroup showed minimal clinical connectivity, consistent with localized fascial restriction and limited symptom generalization. The high-stiffness/moderate-Fc subgroup displayed stronger cross-domain interactions, with PEG and PCS acting as bridge variables between sensory, functional, and emotional domains, suggesting a transitional stage where increasing fascial stiffness begins to influence broader symptom processes. In contrast, the low-stiffness/high-Fc subgroup exhibited the most interconnected network, with sleep, pain interference, and physical function forming a central triad and emotional–sensory variables linked through sleep. This pattern reflects a more globally coordinated symptom state, consistent with early central sensitization. Overall, the uT + Fc model demonstrated a clear biomechanical-to-clinical gradient, where changes in fascial hydration and conductivity corresponded with progressively denser symptom networks.

The model integrating transverse stiffness with shoulder-flexion ROM highlighted how mechanical restriction and functional mobility jointly shape symptom organization. The low-stiffness/limited-ROM subgroup showed moderate connectivity, with Sleep serving as a central hub linking emotional distress, sensory sensitivity, and function. This suggests that movement limitation may coincide with early involvement of fatigue and affective symptoms. The high-stiffness/ ROM subgroup had the most cohesive network, characterized by strong links among PEG, PCS, and Function, and bridge roles for PHQ-2 and PPT. This pattern reflects a stage where individuals maintain functional performance but exhibit heightened cognitive–emotional engagement with pain. In contrast, the flexible, low-to-moderate stiffness subgroup showed a sparse and minimally coupled network, consistent with a lower-symptom or partially recovered state. These findings indicate that joint mobility modifies how stiffness relates to symptom complexity, with greater mechanical–functional mismatch corresponding to stronger multidomain interaction.

Across all three clustering approaches, a consistent pattern emerged: myofascial pain reflects a continuum of mechanical and clinical adaptation rather than a simple stiff– versus–normal distinction. Although Normal, Latent, and Active trigger-point categories were represented across all clusters, their distribution did not fully define subgroup structure, reinforcing that mechanical and functional patterns, not clinical labels alone, drove the observed differentiation.

- The uL-only model captured early, fiber-dominant mechanical differences with variable clinical involvement.
- The uT + Fc model reflected fascial compositional changes that aligned with a clear progression from localized symptoms to broader emotional and sensory coupling.
- The uT + ROM model highlighted how movement capacity interacts with stiffness to shape the degree of symptom integration.

Together, these patterns show that as tissue stiffness, conductivity, and mobility change, symptom networks become increasingly coordinated across pain, emotion, sleep, and sensory domains. This progressive shift, from isolated symptoms to multidomain connectivity, suggests that quantitative imaging and network analysis can serve as objective markers of symptom complexity and stage in myofascial pain syndrome.

### D. Clinical Implications

The multimodal findings clarify how mechanical and fascial changes relate to symptom patterns in MPS. Across the three models, the results suggest a gradual shift from localized mechanical alterations to broader involvement of emotional, sensory, and functional domains.

The uL-only model reflects an early, fiber-dominant stage in which increased longitudinal stiffness represents elevated muscle tone but symptoms remain mostly local or mild. Clinical networks in this stage were variably connected, indicating that elevated fiber tension can exist with limited psychosocial or sensory involvement. Patients in this phase may respond well to treatments targeting muscle tension and improving movement patterns, such as stretching, manual therapy, or posture restoration.

The uT + Fc model highlights a transitional stage characterized by increased transverse stiffness and reduced fascial conductivity. These changes were associated with denser symptom networks involving pain interference, emotional distress, and sleep disturbance. This pattern is consistent with fascial densification and altered hydration, which may contribute to early generalization of symptoms. Interventions here may need to address both tissue quality (hydration, elasticity, fascial mobility) and pain-modulating pathways (relaxation, pacing, breathing techniques).

The uT + ROM model represents a more functionally adaptive or sensitized stage, where stiffness interacts with movement capacity and emotional symptoms. In this phase, when there is loss of ROM, it may reflect compensatory changes in movement patterns in order to preserve functional activity. Some individuals may maintain ROM despite increased stiffness, yet show increased cognitive–emotional engagement with pain. This suggests that closer integration of physical and emotional processes, requiring a multimodal approach combining physical therapy, graded exercise, and cognitive or behavioral strategies, may be needed to support both mobility and coping in order to improve function.

Collectively, these findings support the view that MPS is not a regional problem but a combination of biomechanical, neurophysiological and psychological elements that result in clinical adaptation and maladaptation. The combined imaging and network analysis indicate that identification of these structure-function interactions may help guide stage-specific treatment, emphasizing mechanical release in early phases and integrated physical–psychological approaches when findings warrant.

## V. LIMITATIONS AND FUTURE DIRECTIONS

This study has several limitations. First, the sample size was moderate, which may limit the generalizability of the biomechanical clusters and network patterns observed. Larger cohorts are needed to confirm the stability of these subgroups. Second, all imaging and physiological measurements were obtained from the upper trapezius; therefore, the findings may not extend to other regions commonly affected by myofascial pain. Third, the cross-sectional design prevents conclusions about temporal or causal relationships among mechanical, sensory, and emotional factors. Longitudinal studies are needed to clarify how tissue stiffness and symptom networks evolve over time and with treatment.

Methodologically, further work is required to establish between-day test–retest reliability of rotational SWE under standardized conditions. Similarly, applying longitudinal or dynamic network models could provide insight into how symptom interactions change across clinical states or in response to targeted interventions.

Future research should also examine how the mechanically dominant, transitional, and sensitized subgroups respond to specific therapies. Integrating SWE with bioimpedance and motion-based assessments may support mechanism-informed, personalized rehabilitation strategies. The use of portable bioimpedance devices and wearable motion sensors could further enable continuous monitoring of hydration, and mobility in the real world.

## VI. CONCLUSIONS

This study demonstrated that rotational SWE stiffness parameters, alone or combined with BIS, ROM, and patient-reported outcomes, can identify distinct structural and functional phenotypes of MPS. Rotational SWE demonstrated good-to-excellent reliability. In combination with tissue conductivity or joint mobility, transverse stiffness identified distinct biomechanical subgroups that correspond to meaningful differences in clinical symptoms.

Across SWE-only and multimodal clustering models, network analyses showed that individuals with similar mechanical profiles can exhibit markedly different patterns of interacting symptoms, reflecting varying degrees of mechanical restriction, functional adaptation, and psychosocial involvement. The progression from localized mechanical changes to more integrated pain–emotion– function networks suggests that MPS develops along a continuum of biomechanical and clinical adaptation, rather than as a single uniform condition.

These findings demonstrate the value of a multimodal approach for understanding the interplay between tissue properties and symptom complexity in MPS. By linking measurable changes in stiffness, tissue conductivity and mobility with distinct clinical network patterns, this study provides a framework for objective, mechanism-informed assessment. This approach may support personalized treatment strategies that target the dominant physiological or psychosocial findings.

## Data Availability

The de-identified data supporting the findings of this study are available from the corresponding author upon reasonable request, subject to institutional review board requirements, participant privacy protections, and applicable data-use agreements.

## REFERENCES

[1] R. D. Gerwin, “Classification, epidemiology, and natural history of myofascial pain syndrome,” Current Pain and Headache Reports, vol. 5, no. 5, pp. 412–420, Oct. 2001, doi: 10.1007/s11916-001-0052-8.

[2] J. G. Travell and D. G. Simons, Myofascial Pain and Dysfunction: The Trigger Point Manual, vol. 2. Lippincott Williams & Wilkins, 1992.

[3] J. Dommerholt, T. Hooks, L.-W. Chou, and M. Finnegan, “Myofascial pain and treatment: Editorial.,” J Bodyw Mov Ther, vol. 23, no. 3, pp. 521–531, Jul. 2019, doi: 10.1016/j.jbmt.2019.06.009.

[4] J. P. Shah, N. Thaker, J. Heimur, J. V. Aredo, S. Sikdar, and L. Gerber, “Myofascial Trigger Points Then and Now: A Historical and Scientific Perspective.,” PM R, vol. 7, no. 7, pp. 746–761, Jul. 2015, doi: 10.1016/j.pmrj.2015.01.024.

[5] C. Fernández-de-Las-Peñas and J. Dommerholt, “International Consensus on Diagnostic Criteria and Clinical Considerations of Myofascial Trigger Points: A Delphi Study.,” Pain Med, vol. 19, no. 1, pp. 142–150, Jan. 2018, doi: 10.1093/pm/pnx207.

[6] C. T. Paley et al., “Repeatability of Rotational 3-D Shear Wave Elasticity Imaging Measurements in Skeletal Muscle.,” Ultrasound Med Biol, vol. 49, no. 3, pp. 750–760, Mar. 2023, doi: 10.1016/j.ultrasmedbio.2022.10.012.

[7] A. E. Knight et al., “Full Characterization of in vivo Muscle as an Elastic, Incompressible, Transversely Isotropic Material Using Ultrasonic Rotational 3D Shear Wave Elasticity Imaging.,” IEEE Trans Med Imaging, vol. 41, no. 1, pp. 133–144, Jan. 2022, doi: 10.1109/TMI.2021.3106278.

[8] L. Lacourpaille, F. Hug, K. Bouillard, J.-Y. Hogrel, and A. Nordez, “Supersonic shear imaging provides a reliable measurement of resting muscle shear elastic modulus,” Physiol. Meas., vol. 33, no. 3, p. N19, Feb. 2012, doi: 10.1088/0967-3334/33/3/N19.

[9] J. E. Brandenburg et al., “Quantifying passive muscle stiffness in children with and without cerebral palsy using ultrasound shear wave elastography.,” Dev Med Child Neurol, vol. 58, no. 12, pp. 1288–1294, Dec. 2016, doi: 10.1111/dmcn.13179.

[10] A. Bravo-Sánchez, A. Pablo, L. G, J. F, and A.-V. J, “The Applicability of Shear Wave Elastography to Assess Myotendinous Stiffness of Lower Limbs during an Incremental Isometric Strength Test,” PubMed, 2022, doi: 10.3390/s22208033.

[11] C.-J. Hao et al., “Upper trapezius muscle elasticity in cervical myofascial pain syndrome measured using real-time ultrasound shear-wave elastography,” Quant Imaging Med Surg, vol. 13, no. 8, pp. 5168–5181, Aug. 2023, doi: 10.21037/qims-22-797.

[12] S. F. Eby, P. Song, S. Chen, Q. Chen, J. F. Greenleaf, and K.-N. An, “Validation of Shear Wave Elastography in Skeletal Muscle,” Journal of biomechanics, vol. 46, no. 14, p. 10.1016/j.jbiomech.2013.07.033, Jul. 2013, doi: 10.1016/j.jbiomech.2013.07.033.

[13] J.-L. Gennisson, T. Deffieux, M. Fink, and M. Tanter, “Ultrasound elastography: principles and techniques.,” Diagn Interv Imaging, vol. 94, no. 5, pp. 487–495, May 2013, doi: 10.1016/j.diii.2013.01.022.

[14] J. Wilke, F. Krause, L. Vogt, and W. Banzer, “What Is Evidence-Based About Myofascial Chains: A Systematic Review.,” Arch Phys Med Rehabil, vol. 97, no. 3, pp. 454–461, Mar. 2016, doi: 10.1016/j.apmr.2015.07.023.

[15] A. Stecco, R. Stern, I. Fantoni, R. De Caro, and C. Stecco, “Fascial Disorders: Implications for Treatment.,” PM R, vol. 8, no. 2, pp. 161–168, Feb. 2016, doi: 10.1016/j.pmrj.2015.06.006.

[16] P. Wabel, P. Chamney, U. Moissl, and T. Jirka, “Importance of Whole-Body Bioimpedance Spectroscopy for the Management of Fluid Balance,” Blood Purif, vol. 27, no. 1, pp. 75–80, Jan. 2009, doi: 10.1159/000167013.

[17] S. Mense, “Muscle Pain: Mechanisms and Clinical Significance,” Dtsch Arztebl Int, vol. 105, no. 12, pp. 214–219, Mar. 2008, doi: 10.3238/artzebl.2008.0214.

[18] E. E. Krebs et al., “Development and Initial Validation of the PEG, a Three-item Scale Assessing Pain Intensity and Interference,” J Gen Intern Med, vol. 24, no. 6, pp. 733–738, Jun. 2009, doi: 10.1007/s11606-009-0981-1.

[19] B. D. Darnall et al., “Development and Validation of a Daily Pain Catastrophizing Scale,” J Pain, vol. 18, no. 9, pp. 1139–1149, Sep. 2017, doi: 10.1016/j.jpain.2017.05.003.

[20] M. Rose, J. B. Bjorner, B. Gandek, B. Bruce, J. F. Fries, and J. E. Ware, “The PROMIS Physical Function Item Bank Was Calibrated to a Standardized Metric and Shown to Improve Measurement Efficiency,” J Clin Epidemiol, vol. 67, no. 5, pp. 516–526, May 2014, doi: 10.1016/j.jclinepi.2013.10.024.

[21] L. Yu et al., “Development of Short Forms from the PROMIS Sleep Disturbance and Sleep-Related Impairment Item Banks,” Behav Sleep Med, vol. 10, no. 1, pp. 6–24, Dec. 2011, doi: 10.1080/15402002.2012.636266.

[22] A. Sapra, P. Bhandari, S. Sharma, T. Chanpura, and L. Lopp, “Using Generalized Anxiety Disorder-2 (GAD-2) and GAD-7 in a Primary Care Setting,” Cureus, vol. 12, no. 5, p. e8224, doi: 10.7759/cureus.8224.

[23] K. Kroenke, R. L. Spitzer, and J. B. W. Williams, “The Patient Health Questionnaire-2: validity of a two-item depression screener.,” Med Care, vol. 41, no. 11, pp. 1284–1292, Nov. 2003, doi: 10.1097/01.MLR.0000093487.78664.3C.

[24] S. Malek, E. J. Reinhold, and G. S. Pearce, “The Beighton Score as a measure of generalised joint hypermobility,” Rheumatol Int, vol. 41, no. 10, pp. 1707–1716, 2021, doi: 10.1007/s00296-021-04832-4.

[25] K. K. Chen, P. Rolan, M. R. Hutchinson, and R. M. J. de Zoete, “Reliability of Temporal Summation of Pain in Healthy and Clinical Populations: A Systematic Review and Meta-Analysis,” Eur J Pain, vol. 29, no. 8, p. e70097, Sep. 2025, doi: 10.1002/ejp.70097.

[26] H.-H.-P. Ngo, T. Poulard, J. Brum, and J.-L. Gennisson, “Anisotropy in ultrasound shear wave elastography: An add-on to muscles characterization,” Front Physiol, vol. 13, p. 1000612, Sep. 2022, doi: 10.3389/fphys.2022.1000612.

[27] K. S. Cole and R. H. Cole, “Dispersion and Absorption in Dielectrics I. Alternating Current Characteristics,” J. Chem. Phys., vol. 9, no. 4, pp. 341–351, Apr. 1941, doi: 10.1063/1.1750906.

[28] T. K. Koo and M. Y. Li, “A Guideline of Selecting and Reporting Intraclass Correlation Coefficients for Reliability Research.,” J Chiropr Med, vol. 15, no. 2, pp. 155–163, Jun. 2016, doi: 10.1016/j.jcm.2016.02.012.

[29] B. Jm and A. Dg, “Statistical methods for assessing agreement between two methods of clinical measurement,” PubMed, 1986, Accessed: Dec. 09, 2025. [Online]. Available: https://pubmed.ncbi.nlm.nih.gov/2868172/

[30] L. Kaufman and P. J. Rousseeuw, Finding Groups in Data: An Introduction to Cluster Analysis. John Wiley & Sons, 2009.

[31] P. J. Rousseeuw, “Silhouettes: A graphical aid to the interpretation and validation of cluster analysis,” Journal of Computational and Applied Mathematics, vol. 20, pp. 53–65, Nov. 1987, doi: 10.1016/0377-0427(87)90125-7.

[32] W. H. Kruskal and W. A. Wallis, “Use of Ranks in One-Criterion Variance Analysis,” Journal of the American Statistical Association, vol. 47, no. 260, pp. 583–621, 1952, doi: 10.2307/2280779.

[33] S. L. Lauritzen, Graphical Models. Clarendon Press, 1996.

[34] J. Whittaker, Graphical Models in Applied Multivariate Statistics. Wiley Publishing, 2009.

[35] J. Friedman, T. Hastie, and R. Tibshirani, “Sparse inverse covariance estimation with the graphical lasso,” Biostatistics, vol. 9, no. 3, pp. 432–441, Jul. 2008, doi: 10.1093/biostatistics/kxm045.

[36] S. Epskamp, D. Borsboom, and E. I. Fried, “Estimating psychological networks and their accuracy: A tutorial paper,” Behav Res, vol. 50, no. 1, pp. 195–212, Feb. 2018, doi: 10.3758/s13428-017-0862-1.

[37] R. Foygel and M. Drton, “Extended Bayesian Information Criteria for Gaussian Graphical Models.” Accessed: Apr. 07, 2025. [Online]. Available: https://www.researchgate.net/publication/47860378_Extended_Bayesian_Information_Criteria_for_Gaussian_Graphical_Models

[38] L. C. Freeman, “A Set of Measures of Centrality Based on Betweenness,” Sociometry, vol. 40, no. 1, pp. 35–41, 1977, doi: 10.2307/3033543.

[39] T. Opsahl, F. Agneessens, and J. Skvoretz, “Node centrality in weighted networks: Generalizing degree and shortest paths,” Social Networks, vol. 32, no. 3, pp. 245–251, Jul. 2010, doi: 10.1016/j.socnet.2010.03.006.

[40] D. J. Robinaugh, A. J. Millner, and R. J. McNally, “Identifying highly influential nodes in the complicated grief network,” Journal of Abnormal Psychology, vol. 125, no. 6, pp. 747–757, 2016, doi: 10.1037/abn0000181.

[41] M. A. Green, G. Geng, E. Qin, R. Sinkus, S. C. Gandevia, and L. E. Bilston, “Measuring anisotropic muscle stiffness properties using elastography.,” NMR Biomed, vol. 26, no. 11, pp. 1387–1394, Nov. 2013, doi: 10.1002/nbm.2964.

[42] C. Pirri et al., “Ultrasound Imaging of Thoracolumbar Fascia Thickness: Chronic Non-Specific Lower Back Pain versus Healthy Subjects; A Sign of a ‘Frozen Back’?,” Diagnostics (Basel), vol. 13, no. 8, p. 1436, Apr. 2023, doi: 10.3390/diagnostics13081436.

[43] E. Ertekin, Z. S. Kasar, and F. T. Turkdogan, “Is early diagnosis of myofascial pain syndrome possible with the detection of latent trigger points by shear wave elastography?,” Pol J Radiol, vol. 86, pp. e425–e431, Jul. 2021, doi: 10.5114/pjr.2021.108537.

